# Improving Patient Engagement in Phase 2 Clinical Trials with a Trial-specific Patient Decision Aid (tPDA): A Development and Usability Study

**DOI:** 10.1101/2025.02.25.25322591

**Authors:** Iva Halilaj, Relinde I.Y. Lieverse, Cary J.G. Oberije, Lizza E.L. Hendriks, Charlotte Billiet, Ines Joye, Brice van Eeckhout, Anke Wind, Anshu Ankolekar, Philippe Lambin

## Abstract

**Background:** Making informed decisions about clinical trial participation can be overwhelming for patients due to the complexity of trial information, potential risks and benefits, and the emotional burden of a recent diagnosis. Patient decision aids (PDAs) simplify this process by providing clear information on treatment options, empowering patients to actively participate in shared decision-making (SDM) with their doctors. While PDAs have shown promise in various healthcare contexts, their use in clinical trials, particularly in the form of trial-specific PDAs (tPDAs), remains underutilized.

**Objective:** To address the challenge of patient comprehension of traditional clinical trial materials, we developed a freely accessible, user-friendly tPDA within the context of the ImmunoSABR phase 2 trial. The tPDA aimed to enhance informed decision-making regarding trial participation. The primary endpoint was usability, quantitatively measured by the System Usability Scale (SUS). Secondary endpoints included time spent on the tPDA, patient satisfaction ratings, and participants’ self-reported level of understanding of the trial.

**Methods:** We developed the tPDA following the International Patient Decision Aid Standards (IPDAS) and validated through a structured, three-phase iterative evaluation process. Initial evaluation was performed with 17 computer scientists who had expertise in biomedical applications, ensuring technical robustness. The content and usability were further refined through evaluations involving 10 clinicians and 8 medical students, focusing on clinical accuracy and user-friendliness. Lastly, the tool was tested by six patients eligible for the ImmunoSABR trial, to assess real-world applicability and patient-centered design.

**Results:** Evaluations demonstrated the tPDA’s effectiveness in enhancing informed decision-making, directly addressing our primary endpoint of usability with an overall mean SUS score of 79.4, indicative of good usability. Addressing our secondary endpoints, patients completed the tPDA efficiently, with the majority (four out of six) finishing in under 30 minutes, and all but one within 60 minutes. The mean satisfaction rating was 9.5 out of 10. Qualitative feedback highlighted significant improvements in patients’ understanding of the trial details, reinforcing the tPDA’s role in facilitating better patient engagement and comprehension.

**Conclusion:** Our study demonstrates the feasibility and potential of tPDAs to enhance patient comprehension and engagement in clinical trials. Integrating tPDAs offers a valuable addition to traditional paper-based and verbal communication methods, promoting informed decision-making and patient-centered care.

**Trial registration:** ImmunoSABR Protocol Code: NL67629.068.18; EudraCT: 2018–002583-11; Clinicaltrials.gov: NCT03705403 and NCT04604470; ISRCTN ID: ISRCTN49817477; Date of registration: 03-April-2019.

## Introduction

Cancer treatment has seen significant advancements in recent years, particularly in systemic therapy and radiation oncology [1]. These breakthroughs rely heavily on clinical trials to evaluate new therapies and improve patient outcomes. However, despite the potential benefits, patient enrollment in clinical trials remains disappointingly low [2]. The Institute of Medicine reports that 71% of phase III oncology trials fail to meet their enrollment goals [3,4]. This challenge extends beyond academic trials, with around 80% of industry-sponsored trials also experiencing enrollment delays, potentially leading to substantial financial losses for drug developers [5]. This not only hinders the development of new treatments but also delays the translation of research findings into clinical practice, ultimately impacting the quality of care available to patients [6].

Several factors contribute to this under-enrollment. Limited trial accessibility, lack of physician awareness about relevant trials, and patient misconceptions or difficulties understanding trial information all play a role. Many patients harbor misconceptions about the purpose of clinical trials and may overestimate the potential benefits of investigational treatments [7–9]. A recent meta-analysis including 103 studies showed that over 25% of clinical trial participants did not fully understand the nature of their participation or its voluntary nature [10]. Only around half understood the fundamental concepts of trials, such as the reason for using placebos or the process of randomization. Additionally, widespread misconceptions persist among the general public, with some believing that trial participants might not receive optimal care [11]. The process of recruiting patients into trials faces additional hurdles, including strict advertising regulations, the necessity for comprehensive information packages, and external factors like the COVID-19 pandemic [12]. Even when suitable patients are identified, they often struggle to navigate the lengthy, complex, and technical language commonly found in patient information forms and trial documentation [13].

Patient decision aids (PDAs) are tools that provide unbiased information about diseases and their treatment options, and help elicit patients’ personal values so that they can participate actively in the decision-making process with their clinician [14]. This collaborative process is known as shared decision-making (SDM) and is becoming increasingly relevant in oncology as the complexity of treatment options increases and decisions often involve significant trade-offs between treatment efficacy and quality of life [15]. PDAs can support SDM through improved communication and understanding between patients and clinicians [16–18]. PDAs often include exercises to clarify values and address concerns, which is especially vital for underrepresented groups like the elderly and racial/ethnic minorities who may face unique barriers to active patient participation [19,20]. The benefits of PDAs extend to reducing decisional uncertainty, enhancing comprehension, and fostering patient-centered care [14]. While primarily designed for treatment decisions, PDAs also hold potential as valuable tools for informing patients about clinical trials, supplementing standard patient information.

Despite their potential in supporting an informed decision-making process, little attention has been paid to trial-specific PDAs (tPDAs). A 2015 Cochrane review highlighted this gap, finding limited evidence on tPDA effectiveness due to the scarcity of relevant studies [21]. This review identified only one study with two tPDA randomized controlled trials, yielding inconclusive results regarding knowledge, decisional conflict, and trial participation. While some evidence suggested tPDAs might reduce decisional regret, key outcomes like risk perception and values-based decision-making remained unexplored. The review concluded that current evidence is insufficient to determine the effectiveness of tPDAs for informed consent in clinical trials. While the landscape of tPDA research has evolved since the 2015 review, with several studies demonstrating the positive effects of tPDAs on patient knowledge, reducing decisional regret, and improving values clarification [cite relevant studies], significant knowledge gaps persist, particularly regarding the optimal design and implementation of tPDAs for complex clinical trials involving novel therapies.

Our study addresses this need by developing and evaluating an interactive tPDA for the ImmunoSABR trial, a phase II trial investigating a novel combination therapy (NCT03705403) [22]. This multicenter, randomized, phase II trial across 15 centers in six European countries is investigating the efficacy of combining stereotactic radiotherapy and immunotherapy in prolonging progression-free survival (PFS) while minimizing toxicity and preserving quality of life for patients with non-small-cell lung cancer (NSCLC). Early recruitment challenges revealed the limitations of traditional patient information documents, which often proved difficult to understand and failed to adequately convey the trial design, even with visual aids. Recognizing this, we developed the tPDA as a supplementary tool to enhance patient comprehension and facilitate informed decision-making regarding trial participation [23]. This paper describes our rigorous, multi-stakeholder evaluation process, providing evidence for the tPDA’s potential to enhance patient comprehension, facilitate informed decision-making, and foster active engagement in the clinical trial process.

## Methods

The tPDA’s development and validation followed a structured, multi-phase approach, incorporating feedback from diverse stakeholders: computer scientists with expertise in medical applications, physicians and medical students from hospitals in the Netherlands and Belgium, and Dutch patients with stage 4 NSCLC eligible for the ImmunoSABR trial. Each evaluation phase engaged these groups in distinct ways to ensure a comprehensive assessment of the tPDA. The tPDA was developed and tested locally with a subset of eligible patients, rather than being deployed across all ImmunoSABR trial sites, as a broader rollout would have required additional ethical approvals, validated translations and back translations in various languages, and logistical coordination beyond the scope of this study.

The study was conducted in compliance with the ICH Harmonised Tripartite Guideline for Good Clinical Practice and received ethical approval from the medical ethics committee of Maastricht University Medical Centrum (MUMC) on September 19, 2021 (approval number: NL67629.068.18). All patients participating in the ImmunoSABR trial provided oral and written informed consent after receiving comprehensive information about the study. While informed consent was mandatory for participation in the ImmunoSABR trial, it was not required for using the tPDA web app, which was tested as part of the trial’s broader evaluative processes.

### Recruitment

We used a purposive sampling approach to select participants based on their relevance to the tPDA evaluation. The groups and their roles were as follows:

1. Computer scientists (n=18): Individuals from Maastricht University with expertise in medical applications were selected to assess the tPDA’s technical aspects, user interface, and information visualization.
2. Clinicians and medical Students (n=18): Ten clinicians and eight medical students, recruited from hospitals in the Netherlands and Belgium, evaluated the tPDA’s content accuracy, usability, and comprehensibility from a healthcare professional’s perspective.
3. Potential trial participants (n=6): Dutch patients with stage 4 NSCLC, eligible for the ImmunoSABR trial, participated in the final evaluation phase to assess the tPDA’s usability and comprehensibility from a patient’s perspective. Patients eligible for usability testing were required to meet the same inclusion/exclusion criteria as participants in the ImmunoSABR phase 2 trial (Clinicaltrials.gov: NCT03705403). These criteria were stringent, including limitations on metastatic burden, specific organ function thresholds, and performance status requirements.

### Development of the Trial-specific Patient Decision Aid (tPDA)

The initial tPDA prototype was developed adhering to the International Patient Decision Aid Standards (ipdas.ohri.ca), complemented by physician consultations and a comprehensive literature review. To ensure optimal user-friendliness and accessibility, we designed the tPDA as a Progressive Web App (PWA) [24]. This approach allows the tPDA to function seamlessly across various platforms, including desktop and mobile devices, using any standards-compliant browser. The front-end development leveraged JavaScript, HTML5, CSS3, and a JSON manifest. The PWA’s high-level architecture, illustrated in Figure 1, involves a web server hosting the back-end code (written in Node.js) and the user’s device running the front-end code, including the service worker, manifest, HTML, CSS, and JavaScript components. The tPDA’s development then proceeded through a phased approach, incorporating multiple rounds of evaluation and refinement based on feedback from the diverse stakeholder groups.

**Figure 1:**
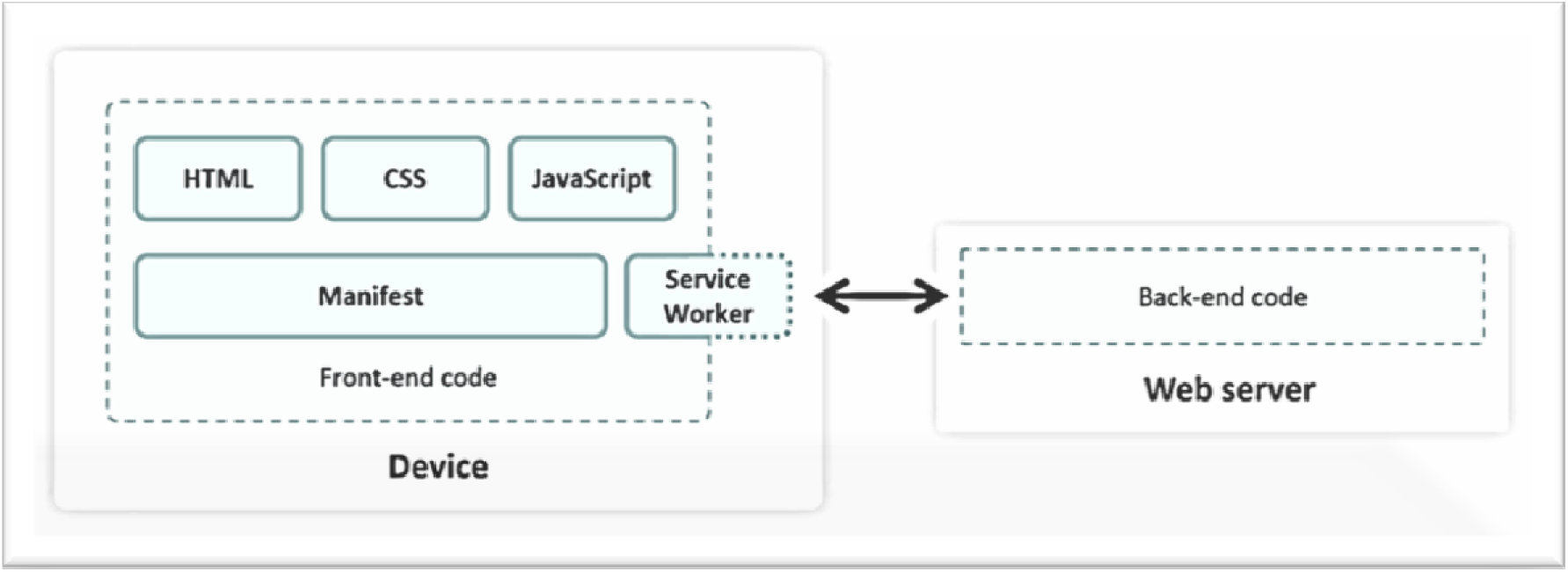
High-level architecture of the Progressive Web App (PWA) used for the tPDA.

### Evaluation instruments

To gather feedback from participants in each round of evaluation, a voluntary questionnaire was introduced. This questionnaire consisted of 17 items, including 10 questions adapted from the validated System Usability Scale (SUS) [25,26]. These questions, translated into Dutch, aimed to evaluate the tPDA’s performance, potential value, and user satisfaction, considering factors like comprehensibility, usability, and the perceived value of the information. The tPDA’s effectiveness and efficiency in supporting decision-making were also assessed. Responses to the SUS questions were recorded on a 5-point Likert scale. The remaining 7 questions were open-ended and designed to gather qualitative feedback on aspects such as the user’s experience with the app, the time taken to complete it, and suggestions for improvement.

### Prototype evaluation

#### Round 1 - Computer scientist review

The initial tPDA prototype was evaluated by 17 computer scientists, focusing on technical functionality, information presentation, and visual design. Their feedback informed the development of the second prototype, which featured improved logical sequencing and enhanced visual representations.

#### Round 2 - Physicians and medical students review

The second tPDA prototype was evaluated by 10 physicians and 8 medical students. Their assessment focused on the accuracy of the information presented, overall usability, and comprehensibility. Feedback from this round was incorporated into the development of the third prototype.

#### Round 3 - Final review and potential trial participants feedback

The final tPDA version underwent evaluation by potential ImmunoSABR trial participants. Their feedback focused on the app’s comprehensibility, usability, and the perceived value of the information presented. This crucial input provided insights into the tPDA’s real-world impact on the decision-making process for potential trial participants.

**Figure 2:**
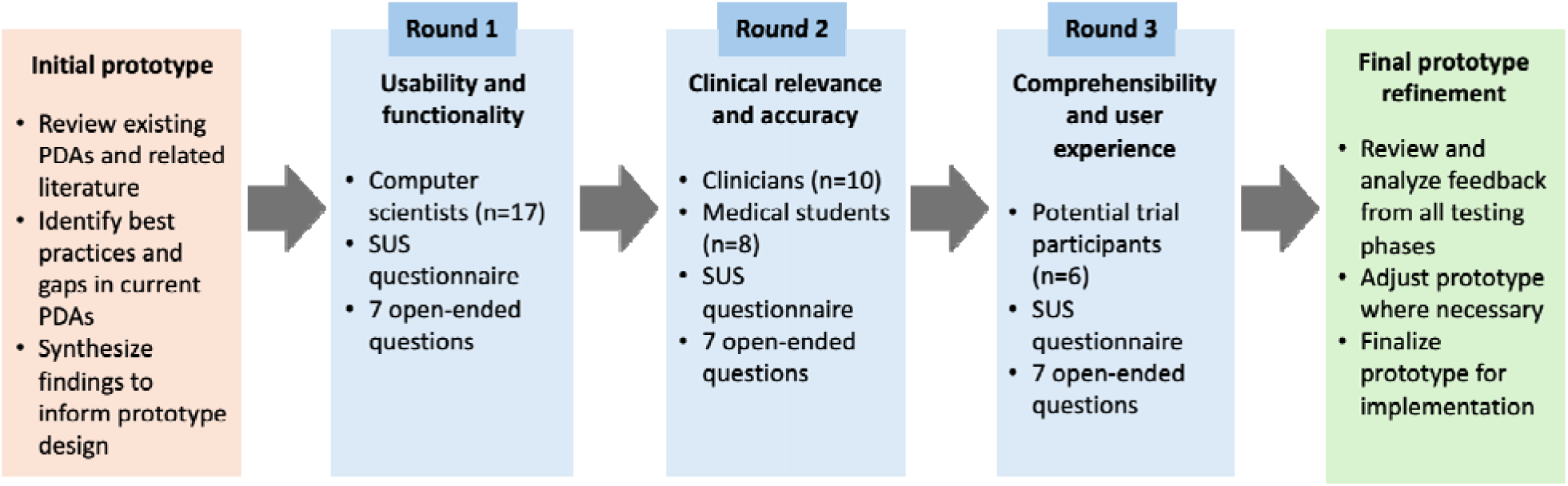
The iterative development and evaluation process of the tPDA, incorporating feedback from computer scientists, clinicians, and potential trial participants.

### Data analysis

Adhering to standardized SUS methodology, raw scores (0-40) were converted to a 0-100 scale and ranked in percentiles for easier interpretation [25]. Given the tPDA’s specific purpose for one-time trial information rather than regular use, we omitted the SUS item “I will use this app frequently.” A recent study indicates that as long as the multiplier is adjusted appropriately (from 2.5 to 2.7), this will not significantly affect the final scores [27]. Data analysis was performed using SPSS 29.0.1. Qualitative feedback collected through open-ended survey responses was used to complement usability metrics. Given the brevity of responses, no formal thematic analysis was conducted, as we did not use structured qualitative data collection methods, such as interviews or focus groups. Instead, responses were reviewed descriptively to identify usability concerns and guide iterative improvements.

## Results

A total of 34 individuals participated in the evaluation of the tPDA across three rounds. This included 17 computer scientists, 10 physicians, 8 medical students, and 6 potential trial participants. The majority of the clinicians were female (77.8%) and aged below 30 years (55.6%). Among the physicians, radiation oncologists constituted the largest group (27.8%), followed by medical oncologists (16.7%). The medical students had less than 10 years of experience and clinicians between 10-20 years of experience. Full demographic and clinical characteristics of the clinicians and medical students are presented in Table 1.

**Table 1:**
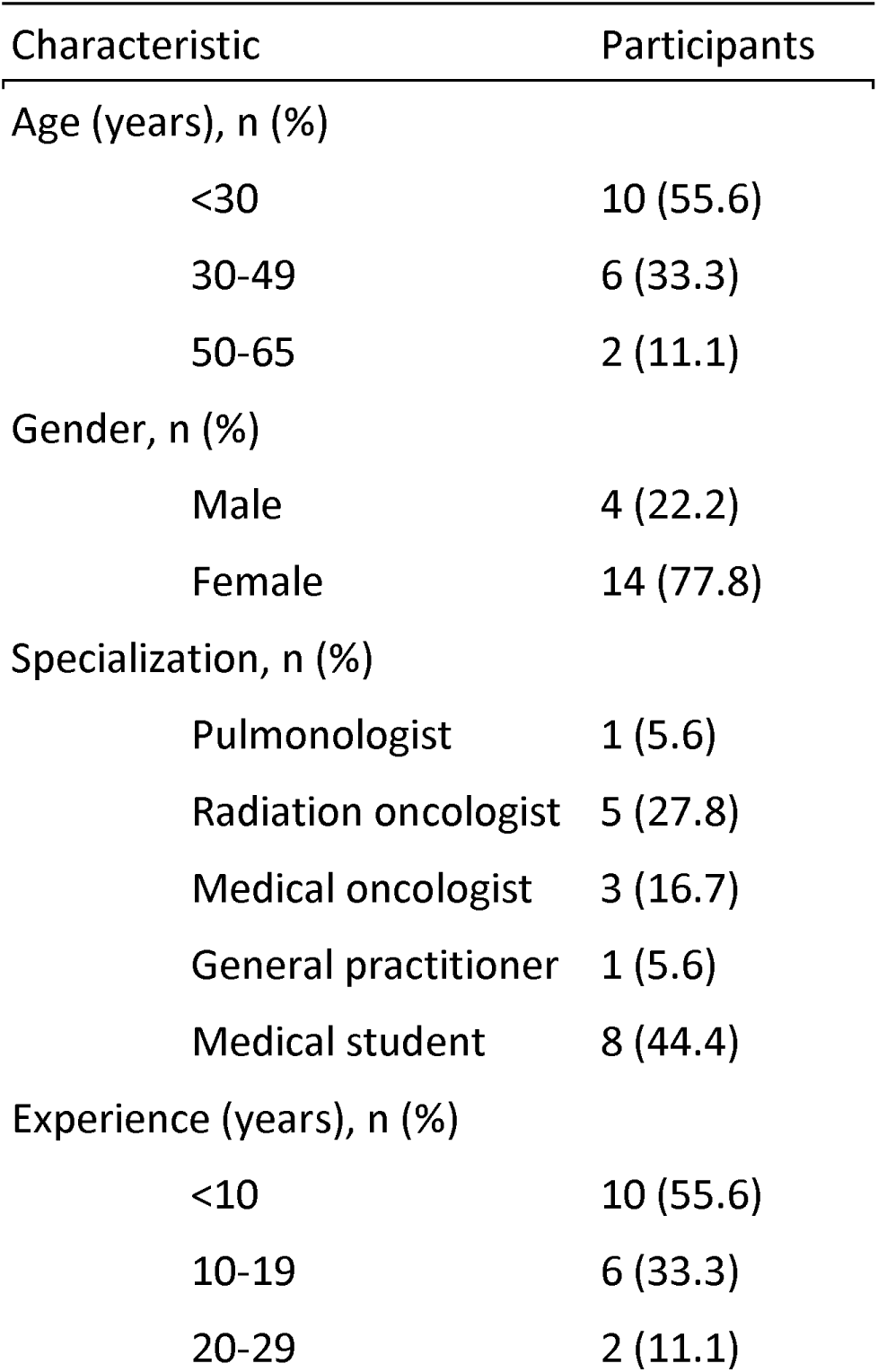
Demographic characteristics of clinician participants (n=18).

### Overall usability assessment and prototype evolution

The tPDA underwent three rounds of iterative evaluation, with each round focusing on specific aspects of the app and incorporating feedback from different stakeholder groups. Across all evaluation rounds and participant groups, the tPDA demonstrated consistently favorable usability, as evidenced by the SUS scores (Figure 3). It is important to note that SUS scores were collected during iterative testing phases, with different stakeholder groups evaluating successive versions of the tPDA. As different groups participated at each stage, these scores reflect independent assessments rather than direct comparisons of usability improvements over time. The final prototype garnered a commendable overall SUS score of 79.4, placing it within the 85-89th percentile range. Notably, patients, the intended end-users, exhibited particularly high satisfaction, with a median SUS score falling in the “excellent” range. Medical students also expressed strong endorsement, with an average SUS score of 94. Physicians (N=10) generally provided a favorable assessment, with an average SUS score of 79 (85-89th percentile). This positive reception was particularly pronounced among radiation oncologists (n=5), who expressed high satisfaction with an SUS score of 80. Medical oncologists (n=3) and the general practitioner (n=1) also provided positive evaluations, with SUS scores of 81 and 80, respectively. In contrast, the pulmonologist (n=1) gave a lower score of 68.

**Figure 3:**
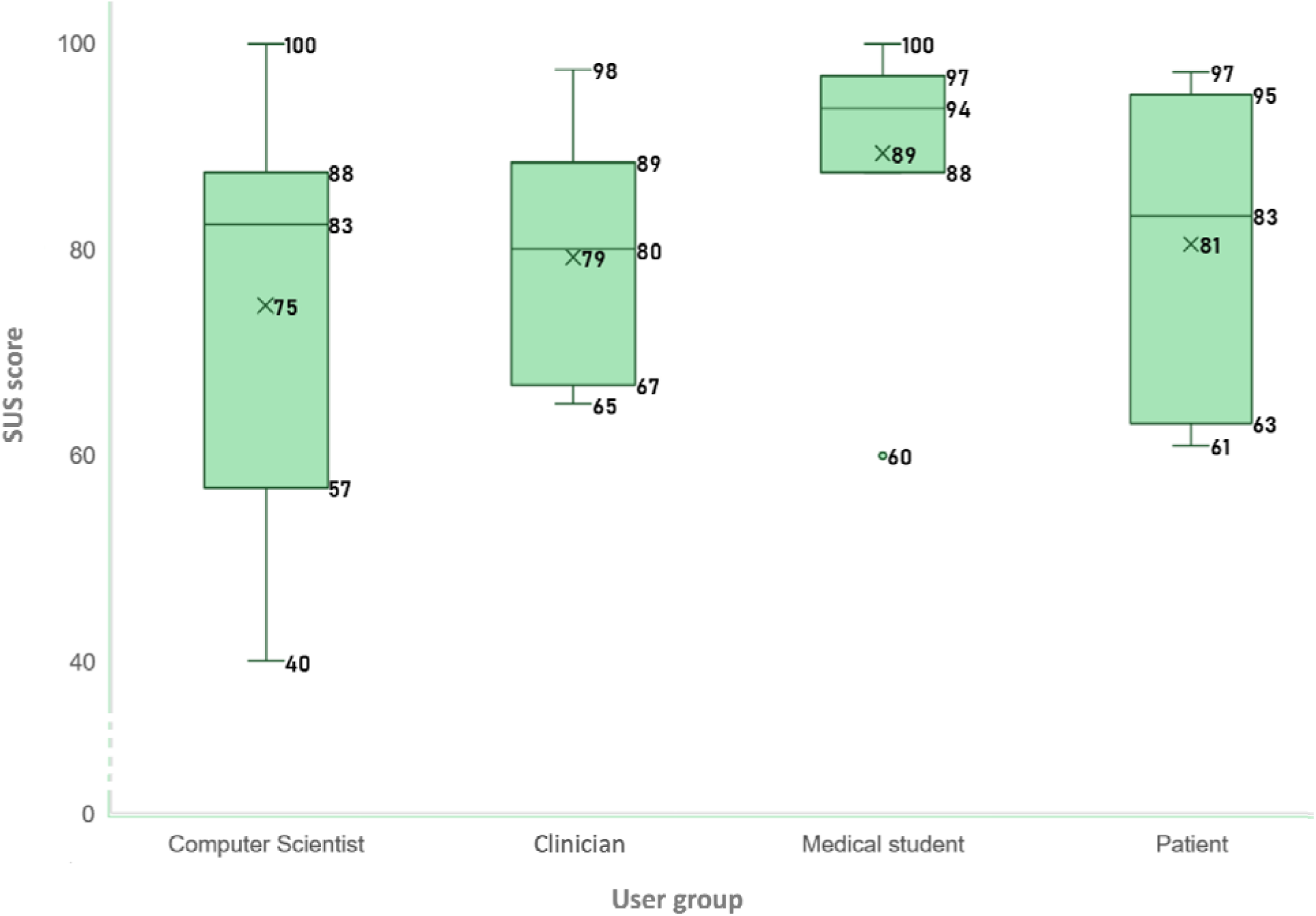
The interquartile range, median and the mean (denoted by ‘x’) of the SUS scores from different participant groups across successive iterations of the tPDA. Scores reflect independent usability assessments rather than a longitudinal comparison.

The iterative refinement process led to significant improvements in the tPDA’s interface and user experience. Figure 4 showcases both the initial prototype and the final version, highlighting the visual and functional enhancements achieved through incorporating feedback from various stakeholders.

**Figure 4:**
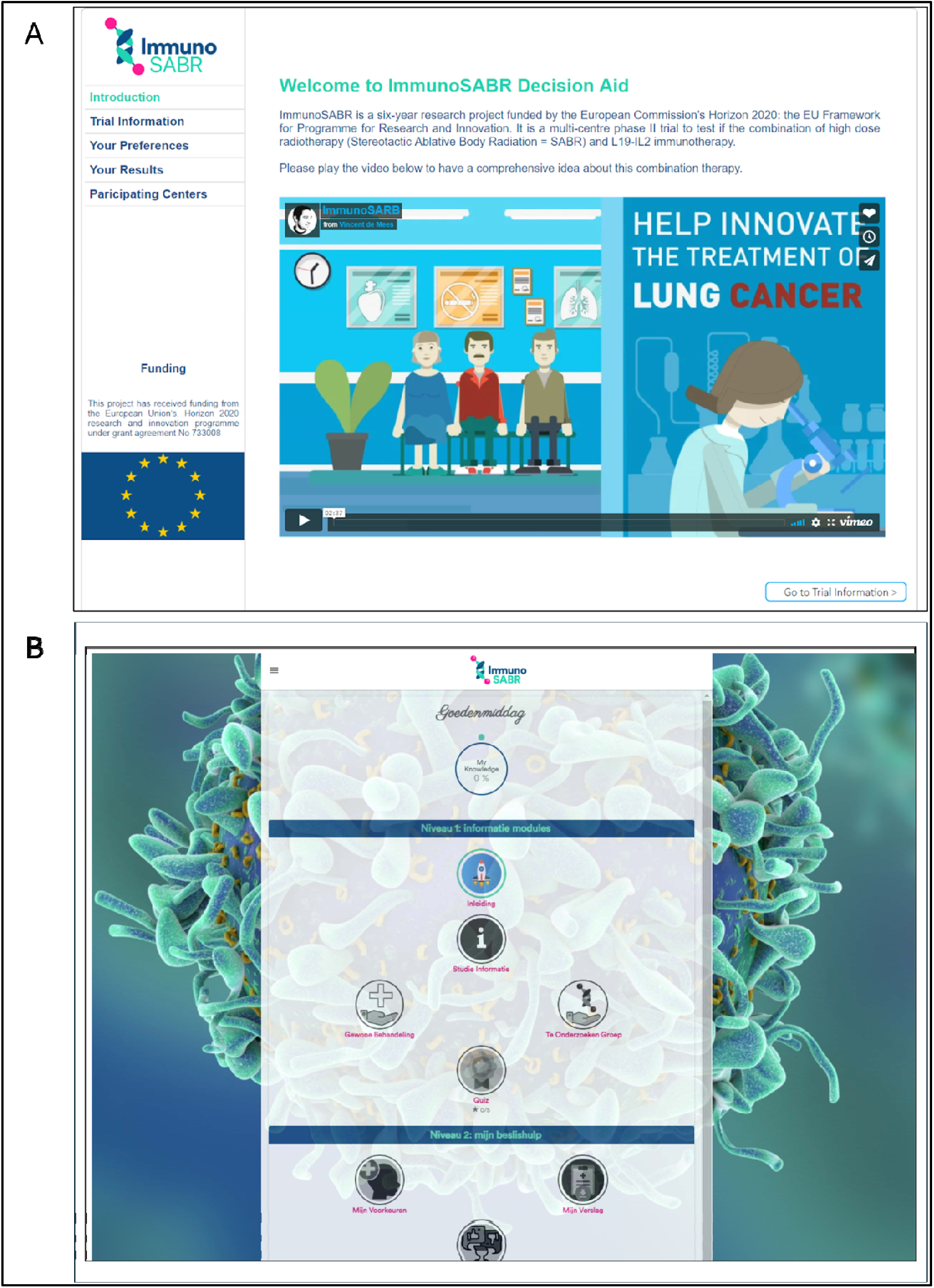
Evolution of the tPDA interface: (A) Initial prototype, and (B) Final prototype after incorporating feedback from multiple evaluation rounds.

### Iterative refinement and feedback

#### Round 1

The initial prototype was first evaluated by 17 computer scientists. Beyond the positive SUS scores, computer scientists provided constructive feedback, highlighting areas for improvement. These included the app’s limited one-time use, cross-device interface inconsistencies, typos and unclear or overlapping text, and the lack of hover features and clear visual cues for clickable elements. These insights guided the development of the second prototype, with a focus on enhancing logical sequencing and visual representations.

#### Round 2

Incorporating feedback from the first round, the second prototype underwent evaluation by 10 physicians and 8 medical students. While the core interface remained largely unchanged, refinements were made to enhance usability and address technical issues. Physicians and medical students provided feedback on several key themes. They advocated for broader language accessibility, suggesting the translation of videos and potentially other content into multiple languages. Additionally, they recommended refining textual content for enhanced clarity and providing further elaboration on specific topics to improve patient understanding. Ensuring accuracy and clarity in animation subtitles was also emphasized. Furthermore, they recommended enhancing the system’s ability to save user input, preventing data loss upon refresh. Specific suggestions, such as incorporating a flowchart illustrating the experimental therapy timeline, aimed to reduce reliance on dense text and improve patient comprehension of complex information. This feedback informed the development of the third prototype, with a focus on addressing these areas and further refining the tPDA’s content and user experience. Following these improvements, the final tPDA prototype was evaluated by potential trial participants.

#### Round 3

The final tPDA prototype was evaluated by six potential ImmunoSABR trial participants. Their feedback was overwhelmingly positive, with an average SUS score of 81, placing it in the 90-95th percentile range. Qualitative feedback revealed that five out of six patients completed the tPDA within 60 minutes, with four finishing in under 30 minutes. The tPDA received high ratings, with four patients giving it a 9 out of 10 and one awarding a perfect 10. Notably, no patients identified any missing topics, indicating that the tPDA adequately covered the necessary information. Patients specifically praised the tPDA’s logical structure, clarity, comprehensibility, and the inclusion of an informative video. Despite the positive feedback, some areas for improvement were also identified. One patient expressed that their interest in trying a new medication was misinterpreted as a potential reason for not participating in the trial, suggesting a need for clearer phrasing in certain questions. Another patient suggested refining two questions to offer more nuanced response options.

## Discussion

While clinical trials are crucial for driving progress in cancer treatment, the process of informing and recruiting eligible patients presents persistent challenges [28,29]. Traditional patient information forms, often dense and filled with technical jargon, can impede comprehension and hinder informed decision-making [30]. This gap in effective communication not only jeopardizes patient safety but also raises ethical concerns regarding the validity of informed consent. Recognizing the need for innovative solutions, our study focused on developing and evaluating the ImmunoSABR tPDA, a digital tool designed to enhance patients’ understanding of the ImmunoSABR trial and help them make an informed choice on whether to participate.

### Principal results

Our study provides promising evidence suggesting that the ImmunoSABR tPDA has the potential to enhance patient comprehension and engagement in the clinical trial decision-making process. The positive feedback from patients, coupled with high SUS scores, indicates the tPDA’s effectiveness in delivering information in a clear, concise, and accessible manner. Patients particularly appreciated the logical structure, visual aids, and interactive elements, which likely facilitated their understanding of complex trial concepts.

Completion time varied among participants, with four out of six finishing within 30 minutes and the remaining two requiring up to 60 minutes. The variability in time required suggests that while the tPDA effectively delivers comprehensive trial information, its length may be a limiting factor in certain clinical settings where patients have restricted availability. Future iterations may explore options to streamline content delivery while preserving key decision-making information. Presenting information in an interactive and visually appealing format could empower patients to take ownership of their healthcare choices, aligning with the broader trend towards patient empowerment and SDM in healthcare [31]. The combination of visual aids and interactive questions to check comprehension within the tPDA actively supports this patient-centered approach to learning and decision-making.

### Limitations

While our study offers valuable insights into the potential of tPDAs in clinical trials, it is essential to acknowledge its limitations. The timing of the tPDA’s development, coinciding with the active patient recruitment phase of the ImmunoSABR trial, limited its immediate impact. By the time the tPDA received ethical approval and was introduced to Dutch trial sites, a significant number of participants had already been enrolled, highlighting a missed opportunity for earlier patient engagement. Another key consideration was the ethical responsibility to ensure that the tPDA was functionally and medically sound before exposing vulnerable patients to its content. Digital health interventions designed for clinical decision-making require rigorous validation to prevent potential misinformation or undue patient burden. To address this, we followed an iterative usability process that prioritized early validation with clinicians and computer scientists before engaging patients in the final testing phase. Future studies should prioritize the concurrent development of tPDAs alongside study protocols, ensuring alignment and timely implementation from the outset.

Secondly, despite proactive efforts to engage trial participants through trial nurses, the response rate to the tPDA was modest. This was in part due to the strict eligibility criteria of the ImmunoSABR trial, which restricted usability testing to a narrow patient population. Additionally, the cognitive and logistical demands placed on trial-eligible patients may have reduced participation rates. Given these limitations, the study prioritized usability testing with clinicians and computer scientists in the earlier development phases to refine the tPDA before patient testing. Future studies could address this by recruiting a broader sample of NSCLC patients, including those who are not actively considering trial participation, to assess the usability of tPDAs in a wider clinical context.

The exact number of participants approached remains undetermined due to a lack of systematic tracking at that stage of the recruitment process. This limitation prevents a precise quantification of the engagement rate and comprehensive analysis of the factors influencing participant responsiveness. Future studies should incorporate structured methods for tracking all recruitment interactions to better understand and enhance engagement strategies. We speculate that many participants may have accessed the tPDA but did not complete the embedded evaluation questionnaire. The tPDA’s privacy-centric design, which avoids storing patient data beyond submitted responses, limits our ability to verify this assumption. Additionally, the absence of a separate informed consent process for tPDA usage might affect the interpretation of its impact on patient engagement and decision-making, as the voluntary nature of its use within the trial setting was not explicitly consented to by participants. Clinic understaffing might also have contributed to limited promotion and support for the tPDA. Future studies could benefit from tracking additional metrics, such as the number of patients informed about the tPDA, actual usage statistics, and incomplete feedback forms. This would provide a more nuanced understanding of patient engagement and identify areas for improvement in tPDA implementation and support.

Finally, it is important to recognize the potential limitations regarding the generalizability of our findings. First, it is important to note that the SUS scores reflect perspectives from diverse user groups, including medical students, clinicians, and patients. While clinicians and students may have evaluated usability based on clinical relevance and interface clarity, patients provided direct end-user feedback. This distinction suggests that perceptions of usability may vary based on professional experience and digital literacy. Future studies should further examine these differences, ensuring that usability assessments prioritize patient-centered evaluations. Second, our focus on a digital tool might inadvertently overlook accessibility issues for patients less familiar with technology. Elderly patients, who constitute a significant portion of the NSCLC population, might face challenges navigating a web-based tPDA [32]. Additionally, socioeconomic disparities can hinder access to digital tools [33]. As tPDAs gain prominence, researchers and developers must proactively address these accessibility barriers to ensure equitable access for all patient populations. This could involve not only exploring alternative formats, such as print or video-based tPDAs, but also adapting the digital tPDA itself to better support users with diverse needs. Potential modifications include simplified navigation, larger fonts, audio narration, and adaptive content presentation based on users’ digital proficiency. Implementing such features could enhance usability and ensure that digital decision aids remain inclusive and accessible to all patients. Additionally, guided sessions where patients use the digital tool with assistance from healthcare professionals, such as doctors or nurses, could provide further support for populations that may face challenges with digital literacy or health comprehension [34,35]. This support is particularly crucial for populations that may face challenges with digital literacy or health comprehension.

### Comparison with prior work

Our evaluation of the ImmunoSABR tPDA contributes to the growing body of evidence supporting the positive impact of tPDAs on informed consent and patient engagement in clinical trials. Previous research has demonstrated the effectiveness of tPDAs in various clinical contexts, highlighting improvements in patient knowledge, reduced decisional regret, and increased willingness to participate in trials [36–45]. These studies, from breast cancer prevention to pediatric oncology, have shown that tPDAs can significantly enhance patient comprehension and facilitate informed decision-making by presenting complex trial information in a clear, concise, and accessible manner.

Building on this body of evidence, our study provides further insights into the usability of tPDAs in the context of a phase II trial involving a novel combination therapy. While we did not directly measure improvements in patient knowledge, feedback highlighted the tPDA’s perceived clarity and ease of use, reinforcing the importance of well-designed digital tools in supporting informed decision-making. These findings echo previous work emphasizing the value of multimedia and interactive components in patient education materials [39–41,46].

Additionally, our evaluation involved a diverse range of stakeholders, including computer scientists, clinicians, and patients, providing a comprehensive assessment of the tPDA’s usability and effectiveness from multiple perspectives. This multi-stakeholder approach aligns with recommendations for developing and evaluating digital health interventions, underscoring the importance of a collaborative process in the design and implementation of effective tPDAs [42].

While our study indicates that participants perceived the tPDA as helpful in structuring trial-related information in a clear and accessible manner, we did not directly measure its effect on patient understanding or decision-making. Further research using validated assessment tools is needed to evaluate the impact of tPDAs on comprehension and actual trial enrollment. Indeed, the influence of these aids on actual trial enrollment remains ambiguous. Some studies have reported that despite enhancing decisional outcomes, tPDAs did not significantly increase participants’ intent to enroll in clinical trials [38,43,44]. We found only one study which observed that patients who used a tPDA enrolled in the trial at a greater rate than the control group who were informed by conventional methods (20.6% versus 9%), however the difference between the two groups was not statistically significant [45]. This suggests that while tPDAs can enhance understanding and facilitate informed decision-making, additional factors must be considered to translate increased understanding into increased trial participation. Future studies could incorporate structured qualitative methods, such as semi-structured interviews or focus groups, to gain deeper insights into patient motivations, decision-making processes, and engagement with tPDAs. This approach could provide a richer understanding of how patients interact with digital tools and inform more patient-centered designs.

Using tPDAs to standardize information delivery across trials and healthcare settings is a key advantage highlighted in our findings and echoed in previous research. However, achieving true standardization requires careful consideration of the balance between providing consistent information and allowing for customization to address the unique aspects of different trials and patient populations. Future research could explore the development of adaptable tPDA templates or frameworks that can be tailored to specific contexts while maintaining core elements of standardization. In doing so, it will be essential to ensure that tPDAs remain patient-centered and address the diverse needs and preferences of different patient groups, as highlighted by studies focusing on specific populations such as rural cancer patients and minority cancer survivors.

While tPDAs offer numerous benefits, their development and implementation require investments in software design, clinical collaboration, and ongoing evaluation. The cost-effectiveness of tPDAs, and indeed PDAs in general, remains an area of ongoing investigation, with some studies suggesting potential long-term cost savings due to improved treatment adherence and reduced decisional regret, while others highlight the need for more robust economic evaluations [45,47,48]. Future research should employ standardized methodologies to assess the economic impact of tPDAs, considering both direct and indirect costs and benefits. Additionally, exploring innovative funding models, such as those proposed for universally accessible tPDA platforms, could help address the financial barriers to widespread implementation.

The scalability of tPDAs, particularly in the context of limited resources and diverse patient populations, presents both opportunities and challenges. While our web-based tPDA offers potential for broad accessibility, it remains crucial to ensure equitable access for all patients, including those with limited digital literacy or socioeconomic disadvantages. This includes translating tPDAs into different languages in international trials. Future research should explore strategies to address these accessibility barriers and investigate the feasibility of implementing tPDAs across different healthcare settings, trial contexts, and patient populations. This could involve developing tPDAs in multiple formats (e.g., web-based, print, video) and evaluating their effectiveness compared to traditional patient information documents in larger, more diverse cohorts, including older adults and individuals from different socioeconomic backgrounds [49,50]. A randomized study or A/B testing approach could provide valuable insights into whether tPDAs lead to measurable improvements in patient comprehension, decision-making confidence, and trial enrollment rates compared to conventional materials.

### Conclusions

Our study highlights the feasibility of implementing the ImmunoSABR tPDA to support patient comprehension and informed decision-making in clinical trials. Usability assessments indicated a favorable user experience, with the tPDA achieving a mean SUS score of 79.4, suggesting ‘Good’ usability. Moreover, the majority of participants (four out of six trial participants) were able to complete the tPDA in under 30 minutes, showcasing its efficiency in delivering essential trial information swiftly. The tPDA also received a high mean satisfaction rating of 9.5 out of 10, further validating its acceptance among potential users (patients and clinicians).

The tPDA offers a patient-centered approach to clinical trial education by addressing the limitations of traditional information provision methods and using interactive visual elements. While it has shown promising results in enhancing immediate patient engagement, further research is needed to fully explore its impact on trial enrollment and long-term patient outcomes. The initial positive evaluation results and alignment with existing literature suggest that tPDAs represent a valuable tool for empowering patients and promoting ethical clinical research practices.

Future efforts should focus on addressing implementation challenges, ensuring accessibility for diverse patient populations, and developing standardized yet adaptable tPDA frameworks to facilitate their wider adoption. This will maximize their benefits for both patients and clinical research, potentially leading to improved trial participation rates and more informed patient consent.

## Data Availability

All data produced in the present study are available upon reasonable request to the authors

## Author contributions

PL initiated the project and secured funding. IH, RL, AW, and AA contributed to data analysis. IH, RL, AW, AA, PL, CO and LH were responsible for drafting and revising the manuscript. All authors read and approved the final manuscript.

## Abbreviations

IPDAS: International Patient Decision Aid Standards
NSCLC: Non-small-cell lung cancer
PDA: Patient decision aid
PWA: Progressive web app
SDM: Shared decision-making
SUS: System Usability Scale
tPDA: Trial patient decision aid

