## Supplementary Tables for "Improving Patient Engagement in Phase 2 Clinical Trials with a Trial-specific Patient Decision Aid (tPDA): A Development and Usability Study"

**Trial registration:** ImmunoSABR Protocol Code: NL67629.068.18; EudraCT: 2018–002583-11; Clinicaltrials.gov: NCT03705403 and NCT04604470; ISRCTN ID: ISRCTN49817477; Date of registration: 03-April-2019.

**Keywords:** patient decision aid; immunotherapy; L19-IL2; clinical trials; usability; informed decision-making; participative medicine

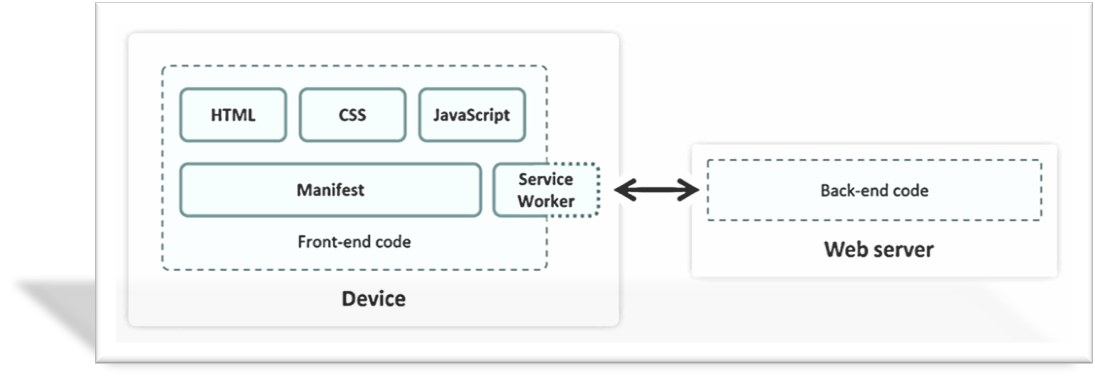

Figure 1: High-level architecture of the Progressive Web App (PWA) used for the tPDA.

#### Evaluation instruments

To gather feedback from participants in each round of evaluation, a voluntary questionnaire was introduced. This questionnaire consisted of 17 items, including 10 questions adapted from the validated System Usability Scale (SUS) [25,26]. These questions, translated into Dutch, aimed to evaluate the tPDA's performance, potential value, and user satisfaction, considering factors like comprehensibility, usability, and the perceived value of the information. The tPDA's effectiveness and efficiency in supporting decision-making were also assessed. Responses to the SUS questions were recorded on a 5-point Likert scale. The remaining 7 questions were open-ended and designed to gather qualitative feedback on aspects such as the user's experience with the app, the time taken to complete it, and suggestions for improvement.

#### Prototype evaluation

##### Round 1 - Computer scientist review

The initial tPDA prototype was evaluated by 17 computer scientists, focusing on technical functionality, information presentation, and visual design. Their feedback informed the development of the second prototype, which featured improved logical sequencing and enhanced visual representations.

##### Round 2 - Physicians and medical students review

The second tPDA prototype was evaluated by 10 physicians and 8 medical students. Their assessment focused on the accuracy of the information presented, overall usability, and comprehensibility. Feedback from this round was incorporated into the development of the third prototype.

##### Round 3 - Final review and potential trial participants feedback

The final tPDA version underwent evaluation by potential ImmunoSABR trial participants. Their feedback focused on the app's comprehensibility, usability, and the perceived value of the information presented. This crucial input provided insights into the tPDA's real-world impact on the decision-making process for potential trial participants.

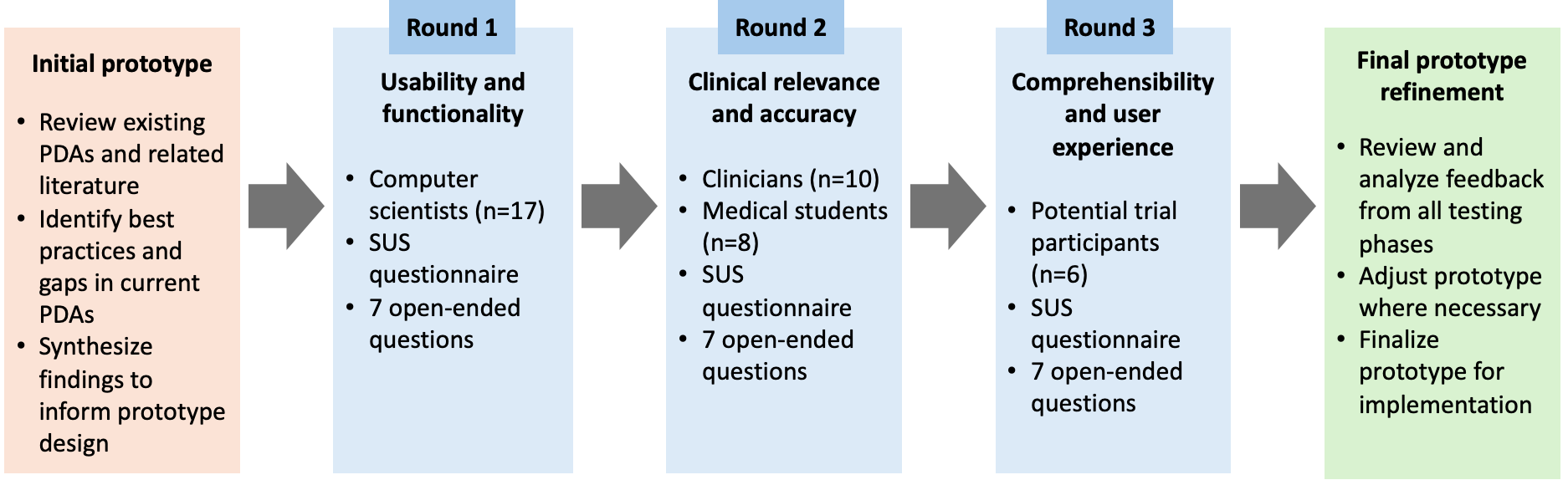

Figure 2: The iterative development and evaluation process of the tPDA, incorporating feedback from computer scientists, clinicians, and potential trial participants.

#### Data analysis

Adhering to standardized SUS methodology, raw scores (0-40) were converted to a 0-100 scale and ranked in percentiles for easier interpretation [25]. Given the tPDA's specific purpose for one-time trial information rather than regular use, we omitted the SUS item "I will use this app frequently." A recent study indicates that as long as the multiplier is adjusted appropriately (from 2.5 to 2.7), this will not significantly affect the final scores [27]. Data analysis was performed using SPSS 29.0.1. Qualitative feedback collected through open-ended survey responses was used to complement usability metrics. Given the brevity of responses, no formal thematic analysis was conducted, as we did not use structured qualitative data collection methods, such as interviews or focus groups. Instead, responses were reviewed descriptively to identify usability concerns and guide iterative improvements.

### Results

A total of 34 individuals participated in the evaluation of the tPDA across three rounds. This included 17 computer scientists, 10 physicians, 8 medical students, and 6 potential trial participants. The majority of the clinicians were female (77.8%) and aged below 30 years (55.6%). Among the physicians, radiation oncologists constituted the largest group (27.8%), followed by medical oncologists (16.7%). The medical students had less than 10 years of experience and clinicians between 10-20 years of experience. Full demographic and clinical characteristics of the clinicians and medical students are presented in Table 1.

Table 1: Demographic characteristics of clinician participants (n=18).

| Characteristic | | Participants |
| --- | --- | --- |
| Age (years), n (%) | | |
|  | <30 | 10 (55.6) |
|  | 30-49 | 6 (33.3) |
|  | 50-65 | 2 (11.1) |
| Gender, n (%) | | |
|  | Male | 4 (22.2) |
|  | Female | 14 (77.8) |
| Specialization, n (%) | | |
|  | Pulmonologist | 1 (5.6) |
|  | Radiation oncologist | 5 (27.8) |
|  | Medical oncologist | 3 (16.7) |
|  | General practitioner | 1 (5.6) |
|  | Medical student | 8 (44.4) |
| Experience (years), n (%) | | |
|  | <10 | 10 (55.6) |
|  | 10-19 | 6 (33.3) |
|  | 20-29 | 2 (11.1) |

#### Overall usability assessment and prototype evolution

The tPDA underwent three rounds of iterative evaluation, with each round focusing on specific aspects of the app and incorporating feedback from different stakeholder groups. Across all evaluation rounds and participant groups, the tPDA demonstrated consistently favorable usability, as evidenced by the SUS scores (Figure 3). It is important to note that SUS scores were collected during iterative testing phases, with different stakeholder groups evaluating successive versions of the tPDA. As different groups participated at each stage, these scores reflect independent assessments rather than direct comparisons of usability improvements over time. The final prototype garnered a commendable overall SUS score of 79.4, placing it within the 85-89th percentile range. Notably, patients, the intended end-users, exhibited particularly high satisfaction, with a median SUS score falling in the "excellent" range. Medical students also expressed strong endorsement, with an average SUS score of 94. Physicians (N=10) generally provided a favorable assessment, with an average SUS score of 79 (85-89th percentile). This positive reception was particularly pronounced among radiation oncologists (n=5), who expressed high satisfaction with an SUS score of 80. Medical oncologists (n=3) and the general practitioner (n=1) also provided positive evaluations, with SUS scores of 81 and 80, respectively. In contrast, the pulmonologist (n=1) gave a lower score of 68.

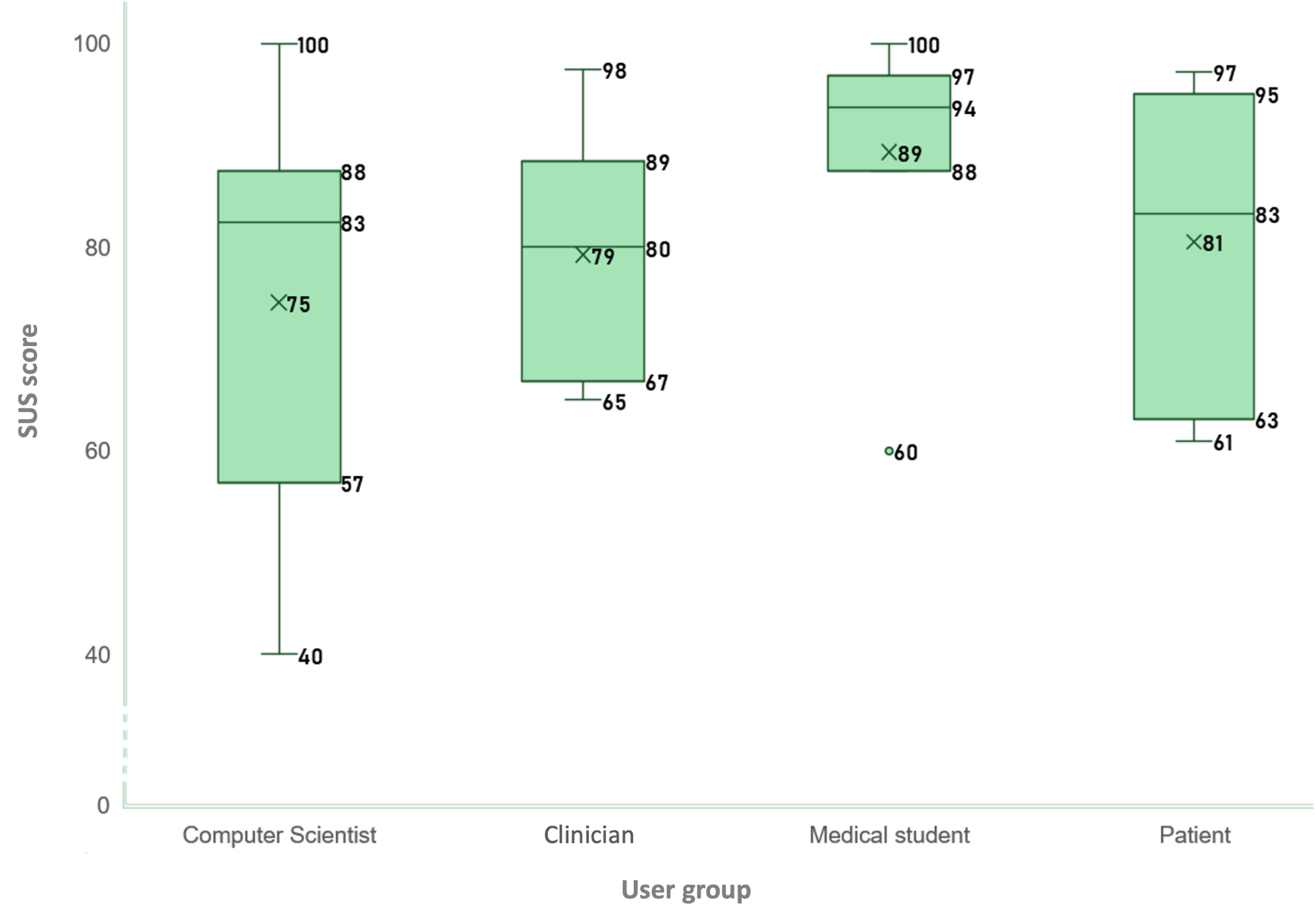

Figure 3: The interquartile range, median and the mean (denoted by ‘x’) of the SUS scores from different participant groups across successive iterations of the tPDA. Scores reflect independent usability assessments rather than a longitudinal comparison.

The iterative refinement process led to significant improvements in the tPDA's interface and user experience. Figure 4 showcases both the initial prototype and the final version, highlighting the visual and functional enhancements achieved through incorporating feedback from various stakeholders.

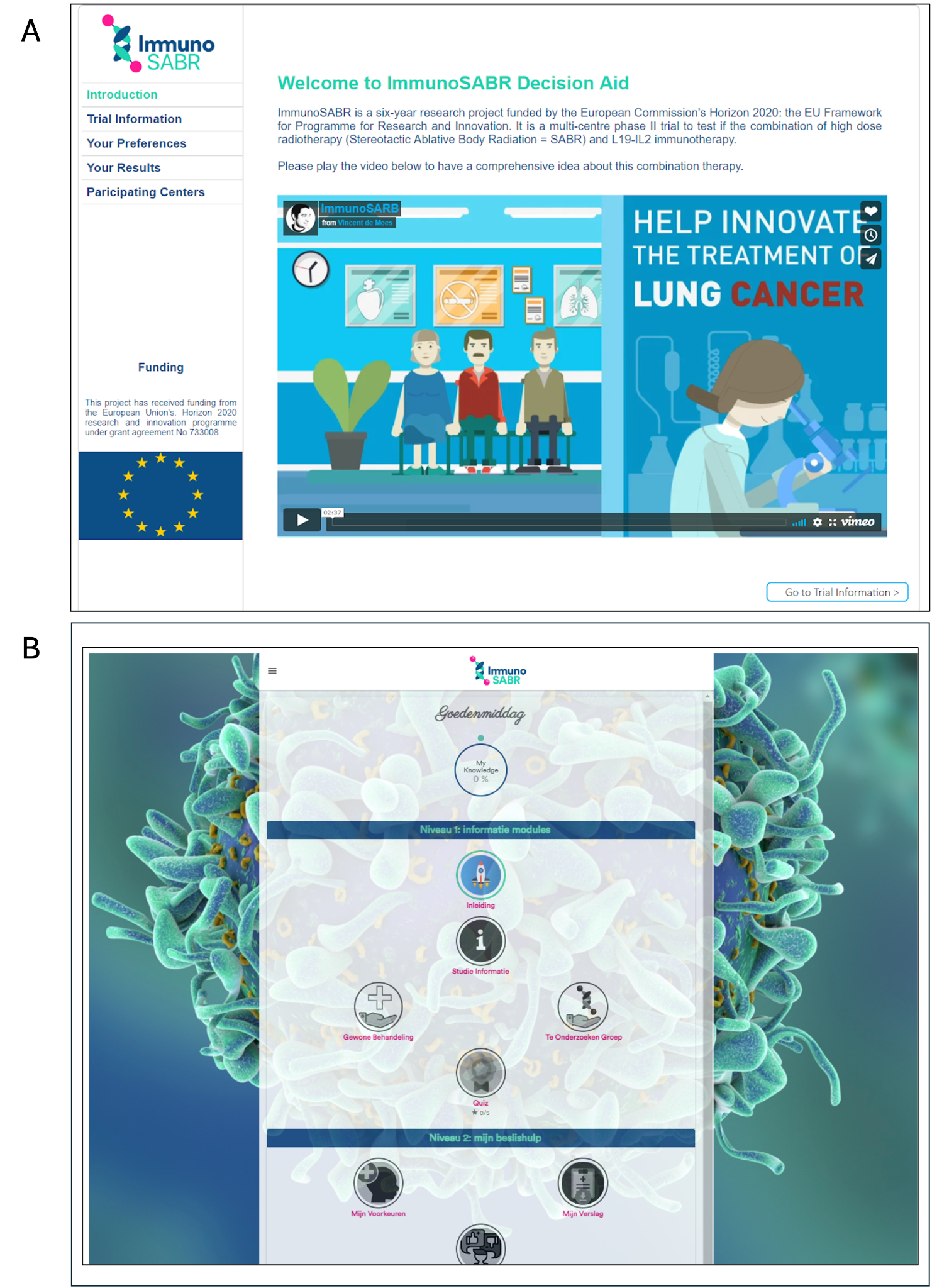

Figure 4: Evolution of the tPDA interface: (A) Initial prototype, and (B) Final prototype after incorporating feedback from multiple evaluation rounds.

#### Iterative refinement and feedback

##### Round 1

The initial prototype was first evaluated by 17 computer scientists. Beyond the positive SUS scores, computer scientists provided constructive feedback, highlighting areas for improvement. These included the app's limited one-time use, cross-device interface inconsistencies, typos and unclear or overlapping text, and the lack of hover features and clear visual cues for clickable elements. These insights guided the development of the second prototype, with a focus on enhancing logical sequencing and visual representations.

### Acknowledgements

#### Funding

Authors acknowledge financial support from ERC advanced grant (ERC-ADG-2015 n° 694812 – Hypoximmuno. Authors also acknowledge financial support from the European Union’s Horizon research and innovation programme under grant agreement: ImmunoSABR n° 733008 (main contributor), CHAIMELEON n° 952172, EuCanImage n° 952103, IMI-OPTIMA n° 101034347, AIDAVA (HORIZON-HLTH-2021-TOOL-06) n°101057062, REALM (HORIZON-HLTH-2022-TOOL-11) n° 101095435, RADIOVAL (HORIZON-HLTH-2021-DISEASE-04-04) n° 101057699, EUCAIM (DIGITAL-2022-CLOUD-AI-02) n° 101100633 and GLIOMATCH n° 101136670.

#### Author contributions

PL initiated the project and secured funding. IH, RL, AW, and AA contributed to data analysis. IH, RL, AW, AA, PL, CO and LH were responsible for drafting and revising the manuscript. All authors read and approved the final manuscript.

### Conflicts of Interest

PL reports, inside the submitted work, board members and minority shares in the company Comunicare Solutions. Disclosures from the last 36 months within and outside the submitted work. Outside of current manuscript: grants/sponsored research agreements from Radiomics SA, Convert Pharmaceuticals SA and LivingMed Biotech srl. He received a presenter fee and/or reimbursement of travel costs/consultancy fee (in cash or in kind) from Astra Zeneca, BHV srl & Roche. PL has/had minority shares in the companies Radiomics SA, Convert pharmaceuticals SA, Comunicare SA, LivingMed Biotech srl and Bactam srl. PL is co-inventor of two issued patents with royalties on radiomics (PCT/NL2014/050248 and PCT/NL2014/050728), licensed to Radiomics SA; one issued patent on mtDNA (PCT/EP2014/059089), licensed to ptTheragnostic/DNAmito; one granted patent on LSRT (PCT/ P126537PC00, US patent No. 12,102,842), licensed to Varian; one issued patent on Radiomic signature of hypoxia (U.S. Patent 11,972,867), licensed to a commercial entity; one issued patent on Prodrugs (WO2019EP64112) without royalties; one non-issued, non-licensed patents on Deep Learning-Radiomics (N2024889) and three non-patented inventions (softwares) licensed to ptTheragnostic/DNAmito, Radiomics SA and Health Innovation Ventures).

LH: none related to the current manuscript, outside of current manuscript: Research funding: Roche Genentech, AstraZeneca, Boehringer Ingelheim, Takeda, Merck, Pfizer, Novartis, Gilead. All to institution. Payment for lectures/presentations/educationals/interviews: MSD, Lilly, Sanofi, GSK, Bayer, high5 oncology. All to institution. Benecke, Medtalks, Medimix, VJOncology. All self. Ad boards: BMS, Lilly, Roche Genentech, Pfizer, Takeda, MSD, Merck, Novartis, Boehringer Ingelheim, Amgen, Janssen, Anheart, Astrazeneca. All to institution. Local PI of clinical trials: AstraZeneca, Novartis, BMS, MSD /Merck, GSK, Takeda, Blueprint Medicines, Roche Genentech, Janssen Pharmaceuticals, Mirati, Abbvie, and Gilead.

BVE reports, inside the submitted work, (minority) shares in the company Comunicare Solutions and being co-inventor of patent PCT/EP2021/053523.

CO: none related to the current manuscript, outside of current manuscript: employee of Kheiron Medical Technologies.

All other authors report no competing interests.

### Abbreviations

IPDAS: International Patient Decision Aid Standards

NSCLC: Non-small-cell lung cancer

PDA: Patient decision aid

PWA: Progressive web app

SDM: Shared decision-making

SUS: System Usability Scale

tPDA: Trial patient decision aid

#### Appendix

Table S1: Overview of questions in the SUS questionnaire.

**SUS questions**

1. I have already received a substantial amount of information regarding the study from both the nursing staff and written materials. This application has assisted me in making a decision regarding my participation in the research.
2. I found the system unnecessarily complex.
3. I thought the system was easy to use.
4. I think that I would need the support of a technical person to be able to use this system.
5. I found the various functions in this system were well integrated.
6. I thought there was too much inconsistency in this system.
7. I would imagine that most people would learn to use this system very quickly.
8. I found the system very cumbersome to use.
9. I felt very confident using the system.
10. I needed to learn a lot of things before I could get going with this system.

**Open questions**

1. Which device did you utilize for the application?
2. How much time did you require to navigate through the application? (<30min; 30-60 min; >60min)
3. On a scale from 1 (poor) to 10 (excellent), how would you rate the application?
4. Is there a subject that you found lacking within the application?
5. Which aspects of the application did you find pleasing?
6. Would you suggest any alterations to the application; if so, what changes would you propose?
7. Do you have any additional comments?

Table S2: Answers to open questions from trial participants

| **Open question** | **Answer** |
| --- | --- |
| Which device did you utilize for the application? | “I had to search for where I could click, as there was no hand icon on the cursor. However, after the first item, that was fine, of course.” |
| How much time did you require to navigate through the application? (<30min; 30-60 min; >60min) | 4x “<30 min”; 1x “30-60 min” |
| On a scale from 1 (poor) to 10 (excellent), how would you rate the application? | 4x “9”; 1x “10” |
| Is there a subject that you found lacking within the application? | 5x “No” |
| Which aspects of the application did you find pleasing? | “The logical sequence of all components.”  “Everything presented clearly in a row.”  “Clear and comprehensible information.”  “It is logically structured.”  “The video.” |
| Would you suggest any alterations to the application; if so, what changes would you propose? | “I responded that I would find testing a new medication exciting, and this emerged as a counterargument in the 'outcome'. However, I disagree with this; while I do find it exciting, it would not be a reason for me not to participate.”  “For Questions 4 and 5, I could only provide an answer with which I was not entirely in agreement. While I am willing to participate in a study involving a new medication, I naturally expect that there is a significant likelihood of benefiting from it. In Question 7, the sentence is incorrect, as a word is missing” |
| Do you have any additional comments? | 5x “No” |

Table S3: Descriptive statistics of the SUS scores for each participant group.

| **Participant group** | **Mean** | **Max** | **Min** | **Std dev** |
| --- | --- | --- | --- | --- |
| Computer Scientist | 83 | 100 | 57 | 18.4 |
| Physicians | 79 | 99 | 65 | 11.8 |
| Medical Students | 94 | 100 | 88 | 12.7 |
| Patients | 81 | 97 | 61 | 13.1 |
